# SARS CoV-2 Serosurvey in Addis Ababa, Ethiopia

**DOI:** 10.1101/2020.06.23.20137521

**Authors:** John H. Kempen, Aida Abashawl, Hilkiah Kinfemichael, Mesfin Nigussie Difabachew, Christopher J. Kempen, Melaku Tesfaye Debele, Abel A. Menkir, Maranatha T. Assefa, Eyob H. Asfaw, Leul B. Habtegabriel, Yohannes Sitotaw Addisie, Eric J. Nilles, Joseph C. Longenecker

## Abstract

The global COVID-19 pandemic caused by SARS CoV-2 is causing both mortality/morbidity and collateral social and economic damage related to public panic and aggressive public policy measures to contain the disease worldwide.(1) The epidemic appears to have taken hold much more slowly in sub-Saharan Africa than most of the world.(2) Antibody testing to evaluate the population proportion previously infected with SARS CoV-2 has the potential to guide public policy, but has not been reported so far for sub-Saharan Africa.

---

Because a large proportion of cases of COVID-19 are asymptomatic and because the elderly are disproportionately affected with severe and symptomatic disease, we hypothesized that a young population might have experienced an epidemic without having recognized it before COVID-19 become widely known. Such a prior epidemic might explain slower growth of the epidemic in sub-Saharan Africa than elsewhere. We hypothesized that a severe influenza-like illness was epidemic in Addis Ababa, Ethiopia around the time COVID-19 was recognized, and might represent a prior epidemic. Because responses taken to contain COVID-19 prevent population-based sampling, and due to the urgency of the situation, we undertook a serological study of approximately 100 persons presenting to a laboratory for other reasons, and an additional group convalescent from the outbreak in November 2019/February 2020, to assess the extent of exposure of the population in Addis Ababa and indirectly assess whether this epidemic may have been attributable to SARS CoV-2.

The study was conducted at International Clinical Laboratories in Addis Ababa, Ethiopia, which offers the Abbott IgG test run on the ARCHITECT platform with approval of the Ethiopian Food and Drug Administration to the general public. This test, which has received the European CE mark and an Emergency Use Authorization from the US Food and Drug Administration, has been found to have 100% sensitivity and 99.9% specificity using the Abbott-determined index value cutoff of 1.4 in an independent study by the University of Washington conducted in Idaho, USA.(3) It has not yet been independently validated in Africa.

Subjects enrolled as part of the two groups met the following eligibility criteria: Age 14 years or higher; resident in Addis Ababa for all of November 2019-February 2020 and no travel outside Ethiopia since November 1, 2019. The following exclusion criteria also were absent: sore throat, runny nose, cough or difficulty breathing and/or hospitalized or quarantined in the last 28 days; measured temperature>99.6 Fahrenheit, resting heart rate >100/minut, and/or resting respiratory rate ≥25/minute); incarceration for a crime; unwilling to participate; or unable to consent. The study was approved in advance by the Institutional Review Boards of MyungSung Medical College (Addis Ababa, Ethiopia) and Partners Healthcare (Boston, USA), and the Addis Ababa Regional Health Bureau; subjects provided written informed consent in Amharic or English.

After training the study team, 99 subjects were recruited from the participating laboratory’s waiting room during May 18-21 inclusive; their characteristics are summarized in the **Table**. Among these, three tested positive for SARS CoV-2 IgG (3.03%, 95% binomial exact confidence interval (CI): 0.63%-8.6%). None of the positive cases were taking medications; one had a chronic runny nose with no other symptoms. Taking into account the sampling scheme and pre-test probability (1.26% based on 1,923 positive results among 152,334 tests as of June 9, 2020(4)), the range of plausible values according to the method of Larremore and colleagues(5) are given in the **Figure**. Forty-five Ethiopians recruited from the investigators’ network who recalled being sick with a COVID-compatible illness between November 1, 2019 and February 29, 2020, were recruited enrolled May 22-27, of whom one tested positive for SARS CoV-2 IgG (2.2%, 95% CI: 0.056%-12%).

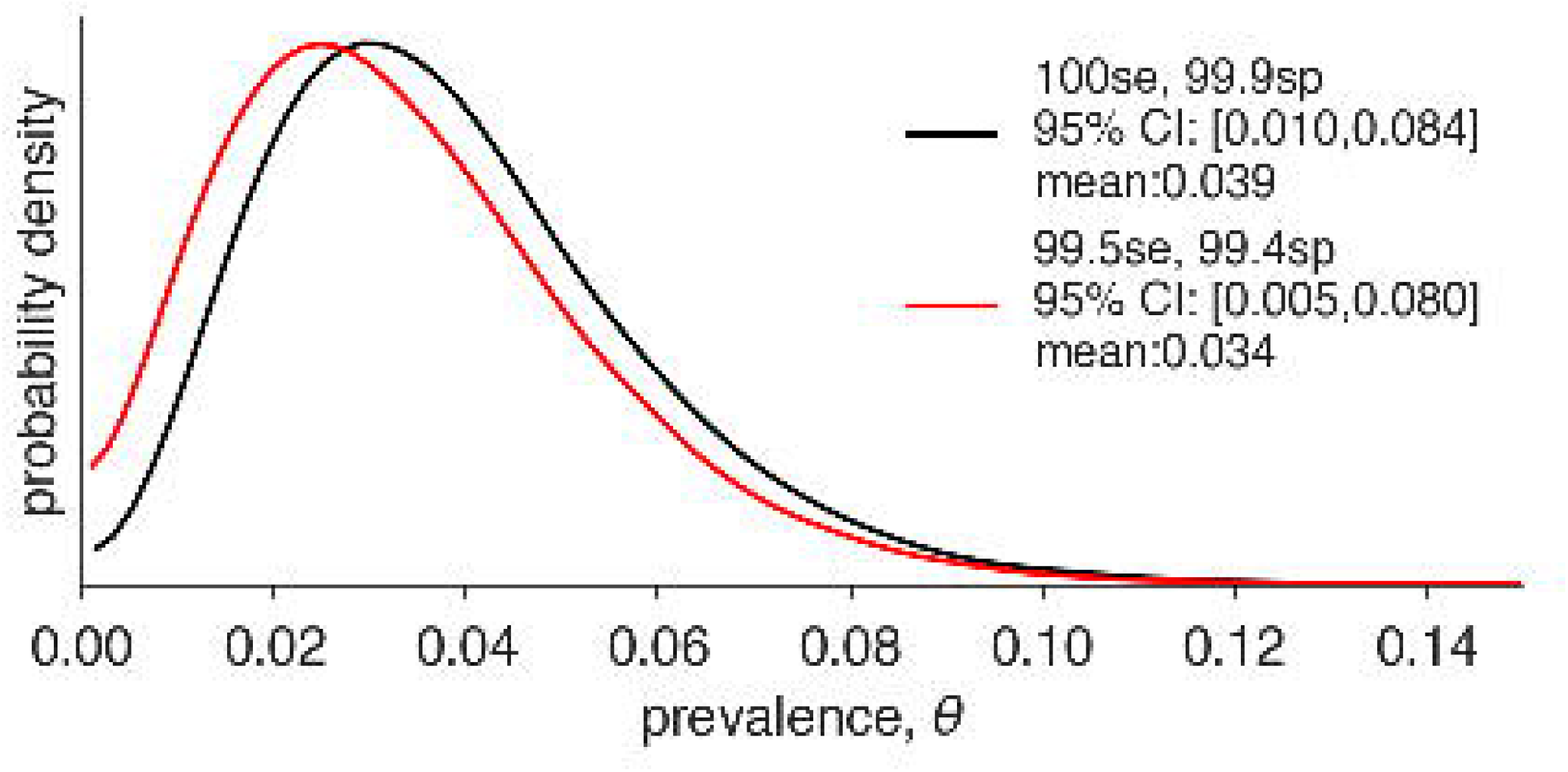
The range of plausible values of SARS CoV-2 IgG seropositivity among asymptomatic persons with no history of COVID-19 infection, expressed as probability density (Addis Ababa, Ethiopia, May 2020), based on sensitivity and specificity results for the test determined by the University of Washington(3) is shown in black. An alternative range under less favorable sensitivity and specificity assumptions is given in red. Se=sensitivity; Sp=specificity.

**Table:** Characteristics of subjects who tested negative or positive for SARS CoV-2 IgG in Addis Ababa, Ethiopia (May, 2020)

|  | SARS CoV-2 IgG |  |
| --- | --- | --- |
|  | Negative | Positive |
| Female | 55% | 67% |
| Age (years), mean | 37.8 | 33.3 |
| Ethiopian | 97% | 100% |
| Total count of people at home, mean | 3.8 | 3.7 |
| Count of children at home, mean | 0.97 | 1.0 |
| Health care worker | 19.4% | 33.3% |
| Hypertension | 12.9% | 0.0% |
| Diabetes mellitus | 12.1% | 0.0% |
| Obesity | 4.4% | 0.0% |
| Lung disease | 2.2% | 0.0% |
| Pregnant (among females) | 9.8% | 0.0% |
| Smoker | 3.3% | 0.0% |
| Any symptoms* | 24.7% | 33.3% |
| Contact with confirmed COVID-19 case | 1.1% | 0.0% |
| Subject thinks (s)he had COVID-19 before** | 1.2% | 0.0% |
| Exposure to traveler*** | 8.6% | 0.0% |
| *Any of the following symptoms each month between November 2019-March 2020 inclusive: fever>37.7 C; feverish (did not measure); chills; headache; coughing or sneezing (more than normal); sore throat; difficulty breathing; runny/stuffy nose; "feeling more tired than usual"; decreased appetite; body or muscle aches; joint pains; decreased sense of smell or taste; nausea; vomiting>2 times; abdominal pain; diarrhea. The subject with positive test result reported a runny/stuffy nose for all five months and no other symptoms. |  |  |
| **Nine subjects were "not sure," including one with positive test result. |  |  |
| ***Family member, friend or co-worker traveled outside Ethiopia between November 2019-March 2020 inclusive. |  |  |

Our results, the first serological general population data on SARS-CoV2 reported from sub-Saharan Africa, are a start toward the vast and important work of characterizing the extent of spread over time in this region with approximately one-seventh of the world’s population. While the results of this kind of sample are limited in scope and not easily generalizable, the results do suggest that the large majority of residents of Addis Ababa have not yet been infected by SARS CoV-2 and are at risk. The results do not suggest any particular risk factors for SARS CoV-2 seropositivity.

Decision-makers in Ethiopia, Africa, and elsewhere all are faced with the dilemma of weighing the trade-offs between the direct consequences of the COVID-19 epidemic and the health, economic and other consequences of aggressive control measures.(2) Given the very young age structure of sub-Saharan Africa with associated lesser direct risk from COVID-19,(6, 7) the risk of famine(8) and limited access to essential services(1) as well as economic problems in this region as collateral problems from COVID-19-associated public panic and restrictive policies, different decisions may be appropriate in the African situation than in countries with an older population age structure and developed economic situation. (9)

## Two Sentence Biography of First Author

Dr. John H. Kempen is Director of Epidemiology for Ophthalmology/Senior Scholar, Massachusetts Eye and Ear/Schepens Eye Research Institute, and Professor of Ophthalmology, Harvard Medical School. He is President of Sight for Souls, a charity developing self-sustaining eye care systems in developing countries, and spends much of the year doing development work, teaching, and research in Addis Ababa, Ethiopia.

## Data Availability

Not available

## Acknowledgments

We thank Daniel B. Larremore, PhD and Yonatan Grad, PhD for preparing the Figure in accordance with their method.(5)

## References

1. Roberton T, Carter ED, Chou VB, Stegmuller AR, Jackson BD, Tam Y, et al. Early estimates of the indirect effects of the COVID-19 pandemic on maternal and child mortality in low-income and middle-income countries: a modelling study. The Lancet Global health. 2020 May 12.

2. Thornton J. Covid-19: Africa’s case numbers are rising rapidly, WHO warns. BMJ. 2020 Jun 15;369:m2394.

3. Bryan A, Pepper G, Wener MH, Fink SL, Morishima C, Chaudhary A, et al. Performance Characteristics of the Abbott Architect SARS-CoV-2 IgG Assay and Seroprevalence in Boise, Idaho. J Clin Microbiol. 2020 May 7.

4. Tadesse L. Notification Note on COVID-19 Situational Update. Ministry of Health Ethiopia; 2020.

5. Larremore DB, Fosdick BK, Bubar KM, Zhang S, Kissler SM, Metcalf CJE, et al. Estimating SARS-CoV-2 seroprevalence and epidemiological parameters with uncertainty from serological surveys. medRxiv. 2020:2020.04.15.20067066.

6. Zheng Z, Peng F, Xu B, Zhao J, Liu H, Peng J, et al. Risk factors of critical & mortal COVID-19 cases: A systematic literature review and meta-analysis. J Infect. 2020 Apr 23.

7. Diop BZ, Ngom M, Pougue Biyong C, Pougue Biyong JN. The relatively young and rural population may limit the spread and severity of COVID-19 in Africa: a modelling study. BMJ Glob Health. 2020 May;5(5).

8. Mukiibi E. COVID-19 and the state of food security in Africa. Agric Human Values. 2020 May 14:1–2.

9. Mehtar S, Preiser W, Lakhe NA, Bousso A, TamFum JM, Kallay O, et al. Limiting the spread of COVID-19 in Africa: one size mitigation strategies do not fit all countries. The Lancet Global health. 2020 Jul;8(7):e881–e3.

